## Supplement for "Reproducible, data-driven characterization of sleep based on brain dynamics and transitions from whole-night fMRI"

### **Supplementary Results**

**Motion parameters with sleep stages.**

Averaged motion across six motion parameters decreased from wake to light sleep to deep sleep at night 2. For example, mean (standard deviation) motion for each sleep stage is as follows, N1: 0.043 (0.37); N2: 0.039 (0.033); N3: 0.035 (0.031); REM: 0.035 (0.032); Wake: 0.057 (0.052).

Similarly, the percentage of timepoints retained after censoring decreased from wake to light sleep to deep sleep at night 2. N1: 91%; N2: 93%; N3: 96%; REM: 89%; Wake 90%.”

**EEG spectral features across HMM states**

We conducted spectral analysis for each TR and calculated the average power spectrum of Cz for each common EEG brainwave—Delta (0.5-4 Hz), Theta (4-8 Hz), Alpha (8-13 Hz), Beta (13-30 Hz), and Gamma (30-100 Hz)—across the 21 HMM states. See **Supplementary Figure 10 and 11** for night 2 and night 1 data, respectively. As expected, we found that N3-related states 8 and 10 had highest Delta power in both nights. In addition, the Deep-N2 module had higher power in Theta and Alpha bands compared to the Light-N2 module.

### **Supplementary Discussion**

**The key similarities and differences between the current study and Stevner et al. (2019).**

Both studies demonstrated that HMM states can be effectively divided into meaningful modules solely based on transition probabilities. Furthermore, both studies indicated that pre-sleep wakefulness differs from post-sleep wakefulness.

However, despite the similar approaches used, key differences in data acquisition and analysis make it challenging to directly compare HMM states between these two studies. Firstly, Stevner et al. (2019) collected only 1-hour-long sleep data from 18 participants, whereas our current study includes 8-hour-long sleep data from 12 participants for two consecutive nights. As discussed in the main text, full sleep cycling cannot be obtained from 1-hour long sleep due to the lack of REM stage and incomplete sleep cycles. Secondly, in Stevner et al. (2019) (Figure 4e), the four wake-NREM stages had roughly the same duration. In contrast, in our current study (Night 2, Figure 2A), the N2 stage comprises 43% of total sleep, which aligns with the natural N2 composition of nocturnal sleep stages. This discrepancy might explain the different number of N2-related states found in the two studies, with 3 out of 19 in Stevner et al. (2019) versus 13 out of 21 in our current study.

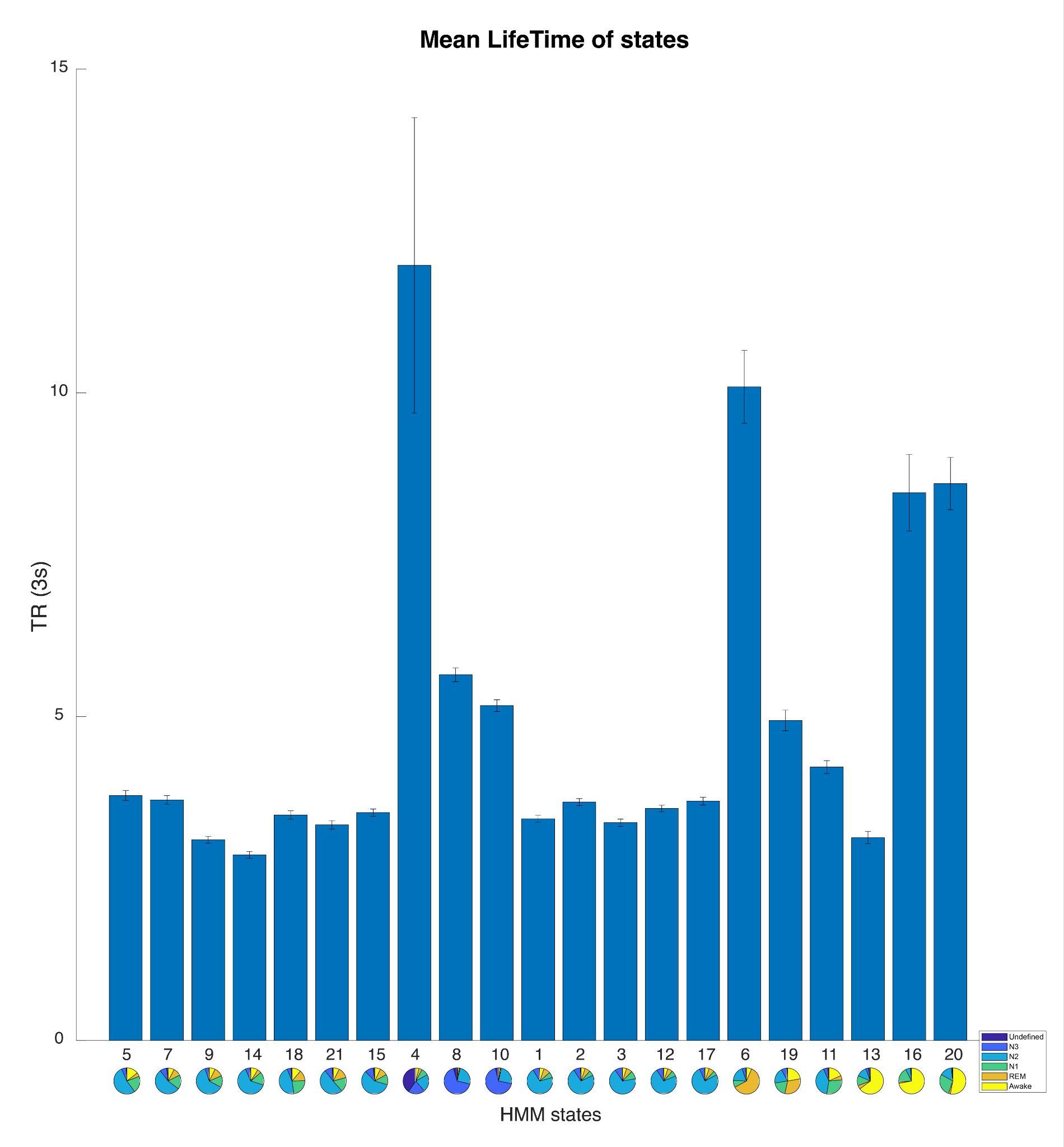

**Supplementary Figure 1.** The mean Lifetime of 21 HMM states. The HMM states are organized based on the results of modular analysis. The error bars represent the standard error of the mean.

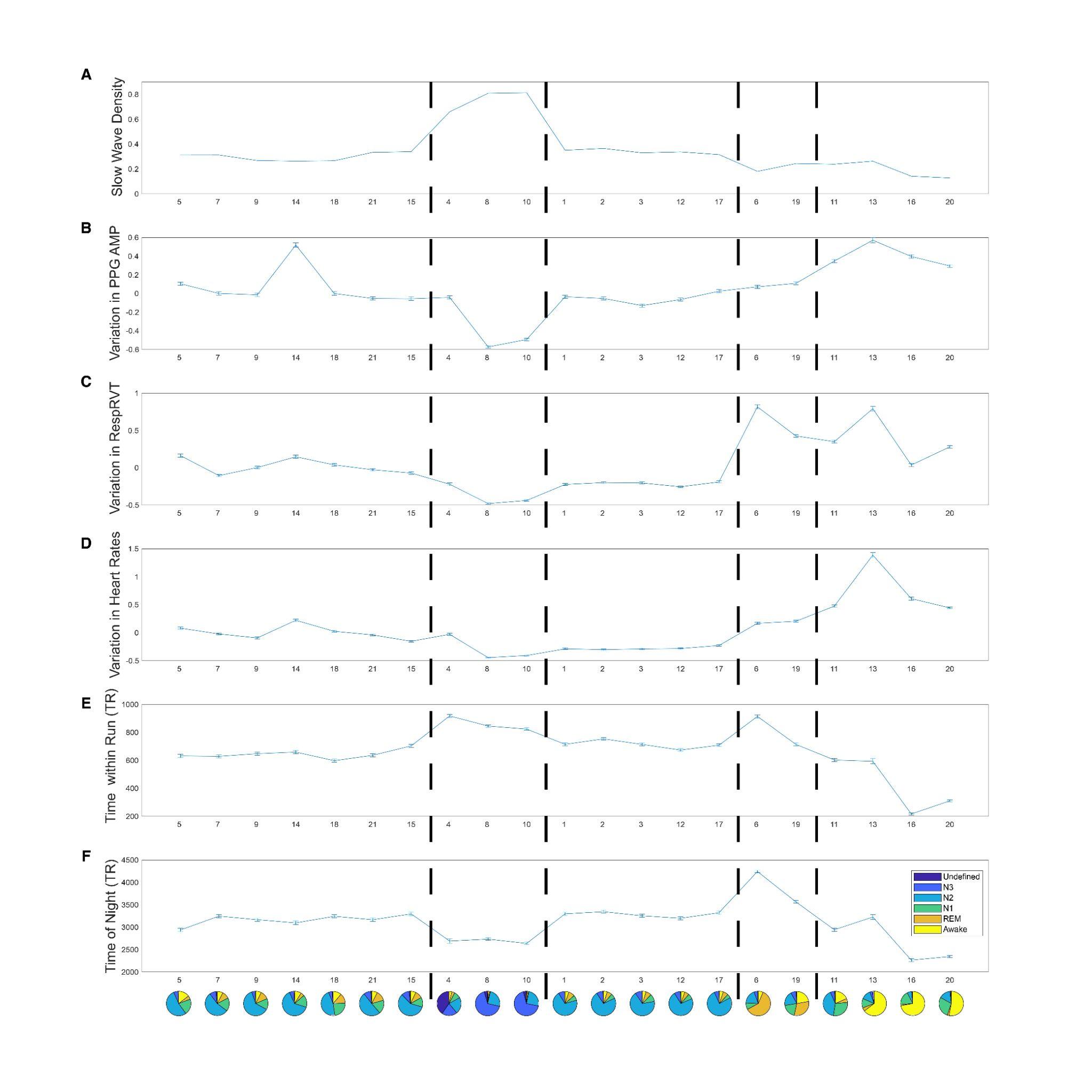

**Supplementary Figure 2.** Physiological variables associated with each HMM state during Night 2. The error bars represent the standard error of the mean. Panel **A**: slow wave density (the percentage of TR that had slow waves); Panel **B**: variation in PPG amplitude (z-score); Panel **C**: variation in Respiratory Volume per Time (z-score); Panel **D**: variation in Heart Rates; Panel **E**: time within a fMRI Run (TR, 3-second); Panel **F**: time since experiment start (TR, 3-second).

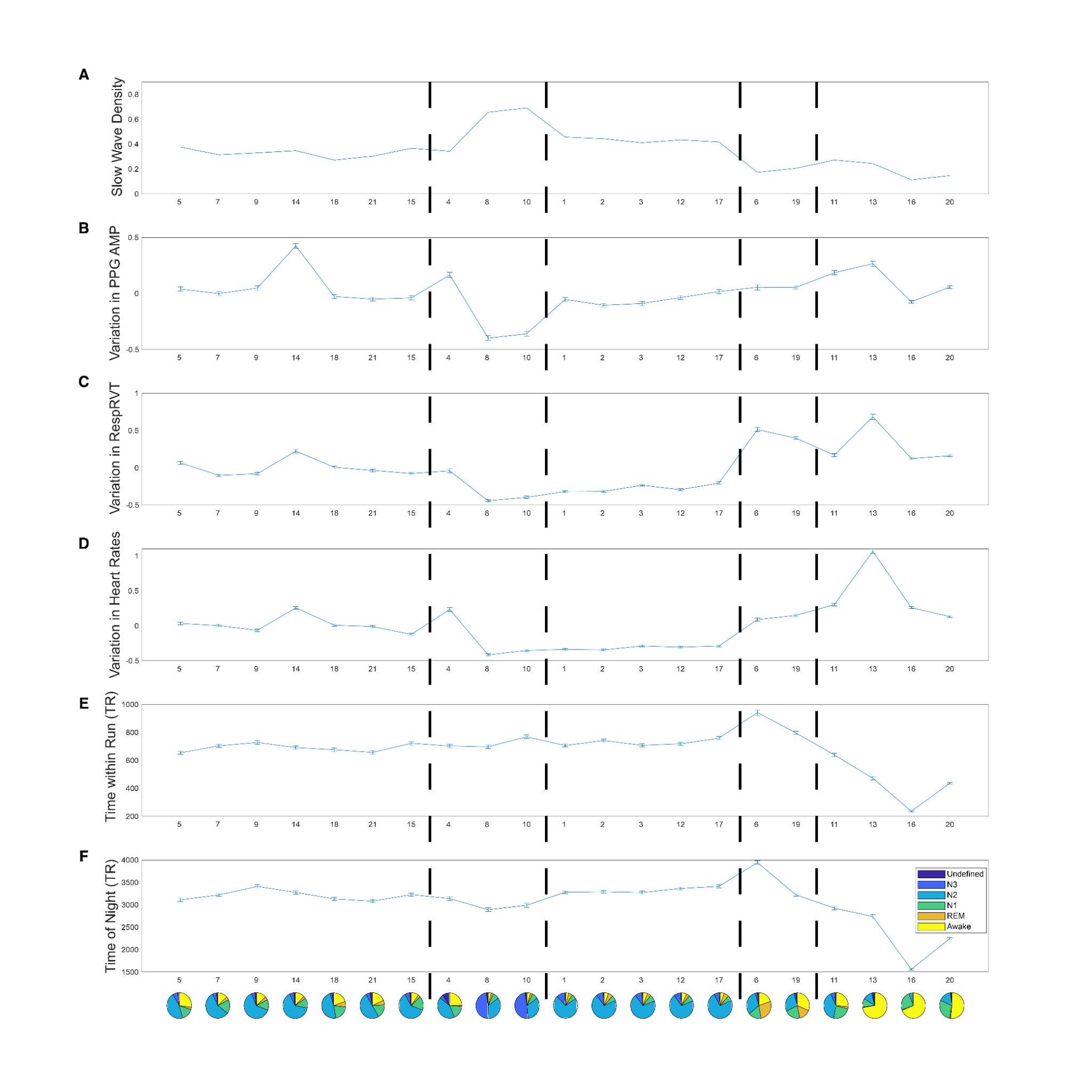

**Supplementary Figure 3.** Physiological variables associated with each HMM state during Night 1. The error bars represent the standard error of the mean. Panel **A**: slow wave density (the percentage of TR that had slow waves); Panel **B**: variation in PPG amplitude (z-score); Panel **C**: variation in Respiratory Volume per Time (z-score); Panel **D**: variation in Heart Rates; Panel **E**: time within a fMRI Run (TR, 3-second); Panel **F**: time since experiment start (TR, 3-second).

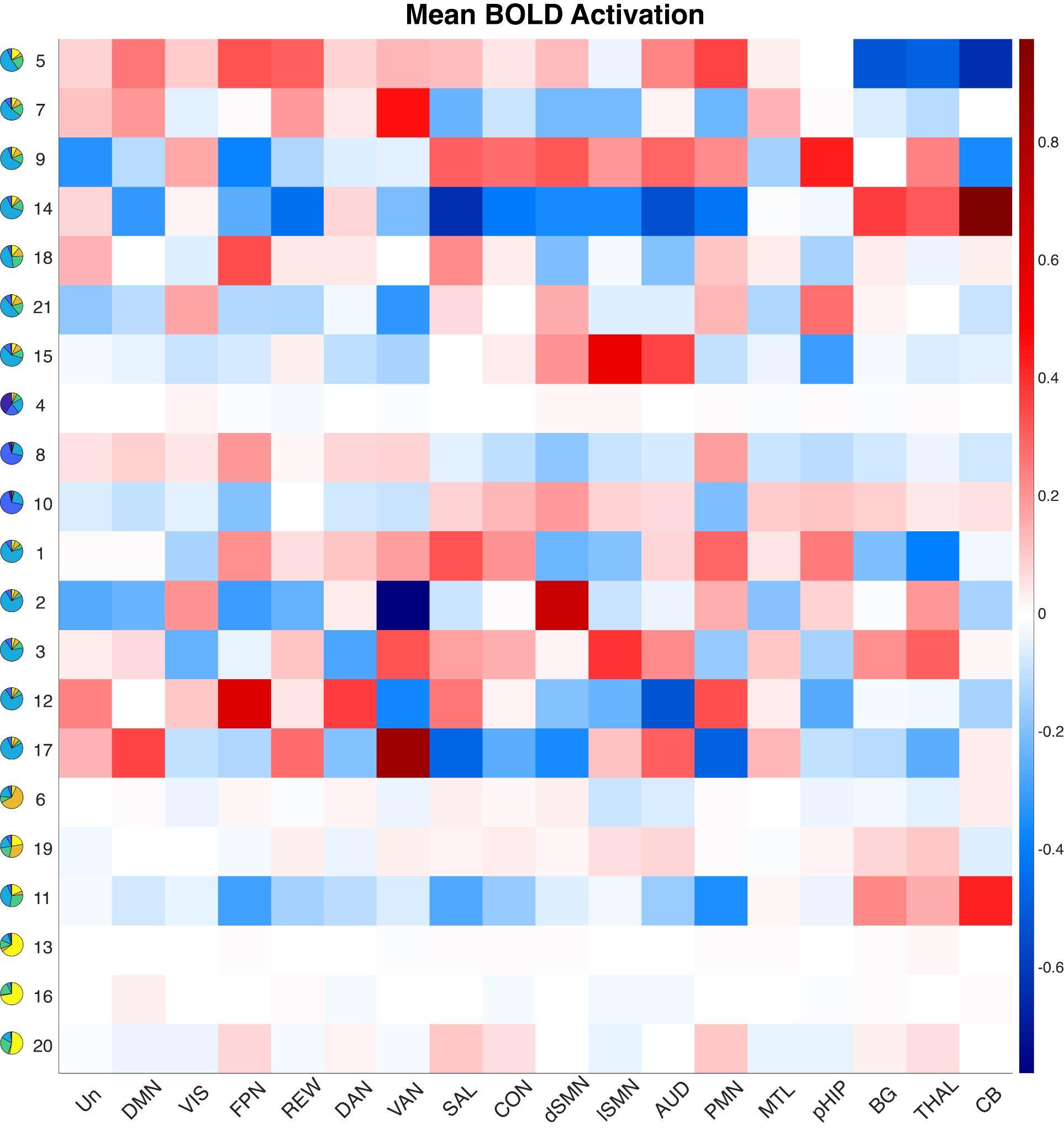

**Supplementary Figure 4.** Mean fMRI activation (percent signal change) for each state relative to baseline averaged over all HMM states. Notes: Un: Undefined Network; DMN: Default Mode Network; VIS: Visual Network; FPN: Frontoparietal Network; REW: Reward Network; DAN: Dorsal Attention Network; VAN: Ventral Attention Network; SAL: Salience Network; CON: Cingulo-Opercular Network; dSMN: Somatomotor Dorsal Network; lSMN: Somatomotor Lateral Network; AUD: Auditory Network; PMN: ParietoMedial Network; MTL: Medial Temporal Network; pHIP: Posterior Hippocampus; BG: Basal Ganglia; THAL: Thalamus; CB: Cerebellar Cortex.

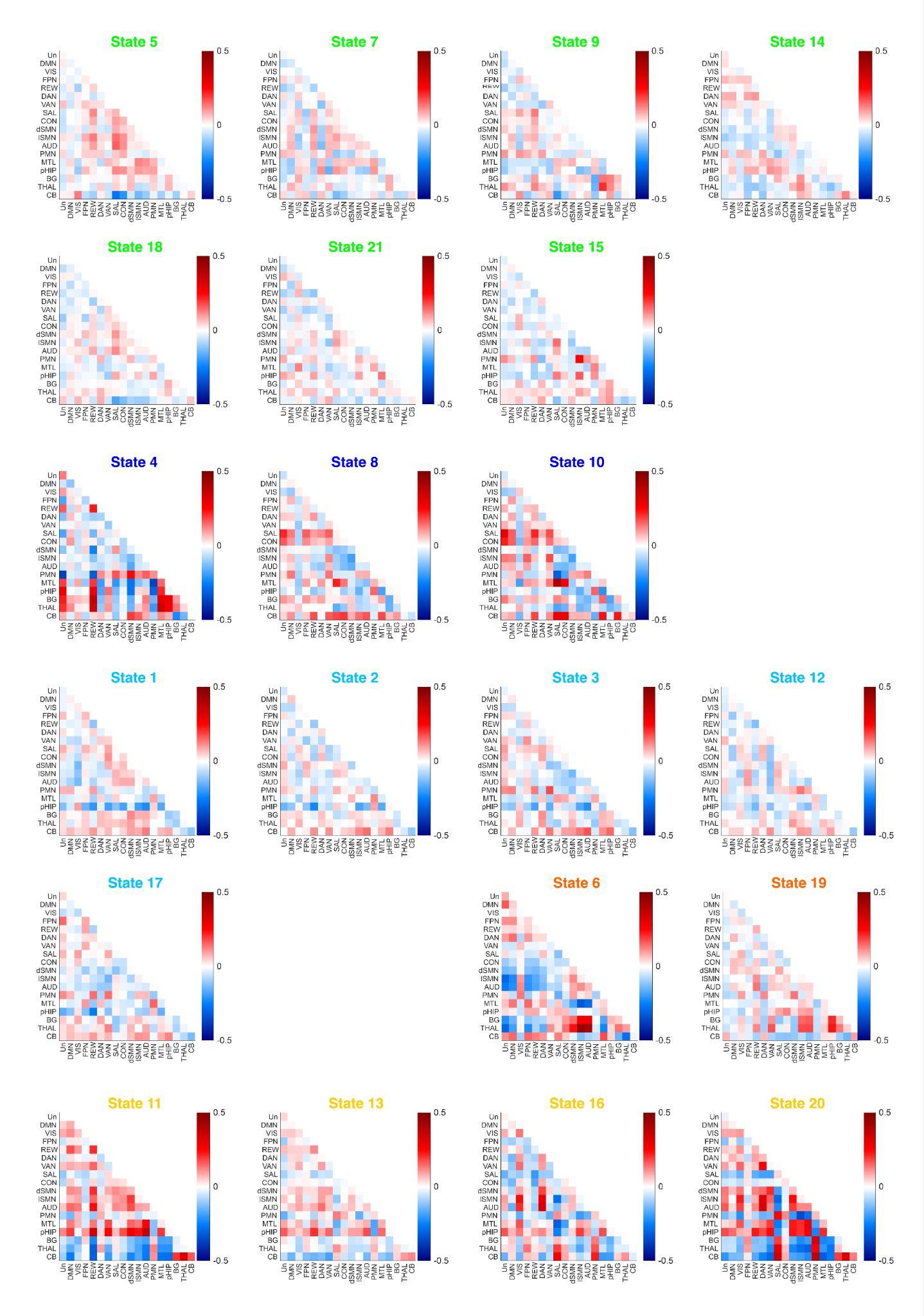

**Supplementary Figure 5.** FC Patterns for each state relative to baseline averaged over all HMM states. HMM states were color-coded based on modules (Green: light N2 module; Dark Blue: N3 module; Light Blue: deep N2 module; Orange: REM module; Yellow: Wake Module). Notes: Y-axis from top to bottom or X-axis from left to right: Un: Undefined Network; DMN: Default Mode Network; VIS: Visual Network; FPN: Frontoparietal Network; REW: Reward Network; DAN: Dorsal Attention Network; VAN: Ventral Attention Network; SAL: Salience Network; CON: Cingulo-Opercular Network; dSMN: Somatomotor Dorsal Network; lSMN: Somatomotor Lateral Network; AUD: Auditory Network; PMN: ParietoMedial Network; MTL: Medial Temporal Network; pHIP: Posterior Hippocampus; BG: Basal Ganglia; THAL: Thalamus; CB: Cerebellar Cortex.

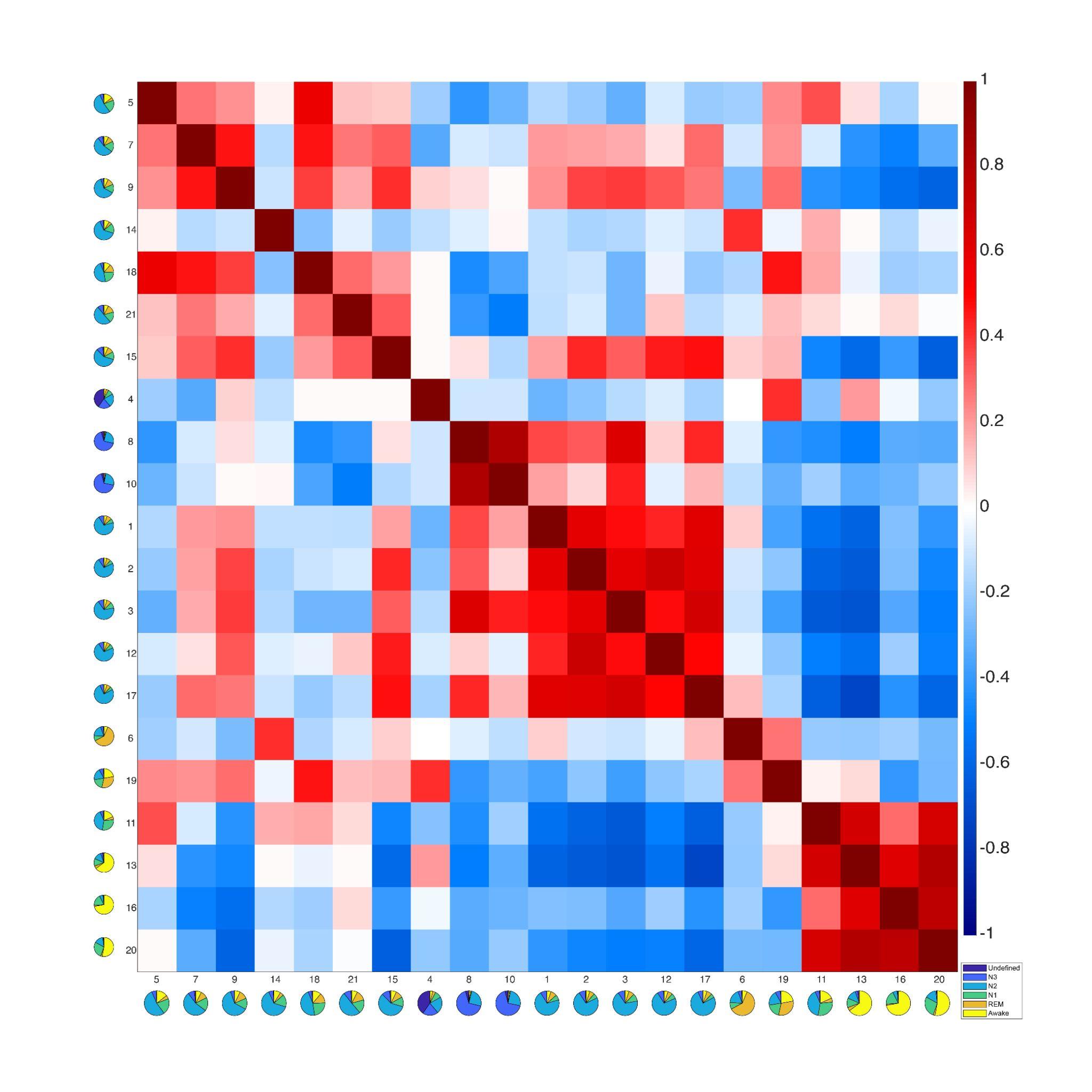

**Supplementary Figure 6.** Correlation matrix between FC patterns of each pair of HMM states. The color bar represents Pearson correlation coefficients. The figure depicts similar modules as the results of the modular analysis in **Figure 3**. For example, states within the Deep-N2 module (states 1, 2, 3, 12, and 17) are highly correlated to each other. These states also show a higher similarity with the N3 module compared to the states within the Light-N2 module (states 5, 7, 9, 18, 21, and 15, except for state 14, which might be due to high physiological variation associated with state 14, see **Supplementary Figure 2 B&D**). REM states 6 and 19 correlated with each other. States within the Wake module (11, 13, 16, and 20) highly correlated with each other.

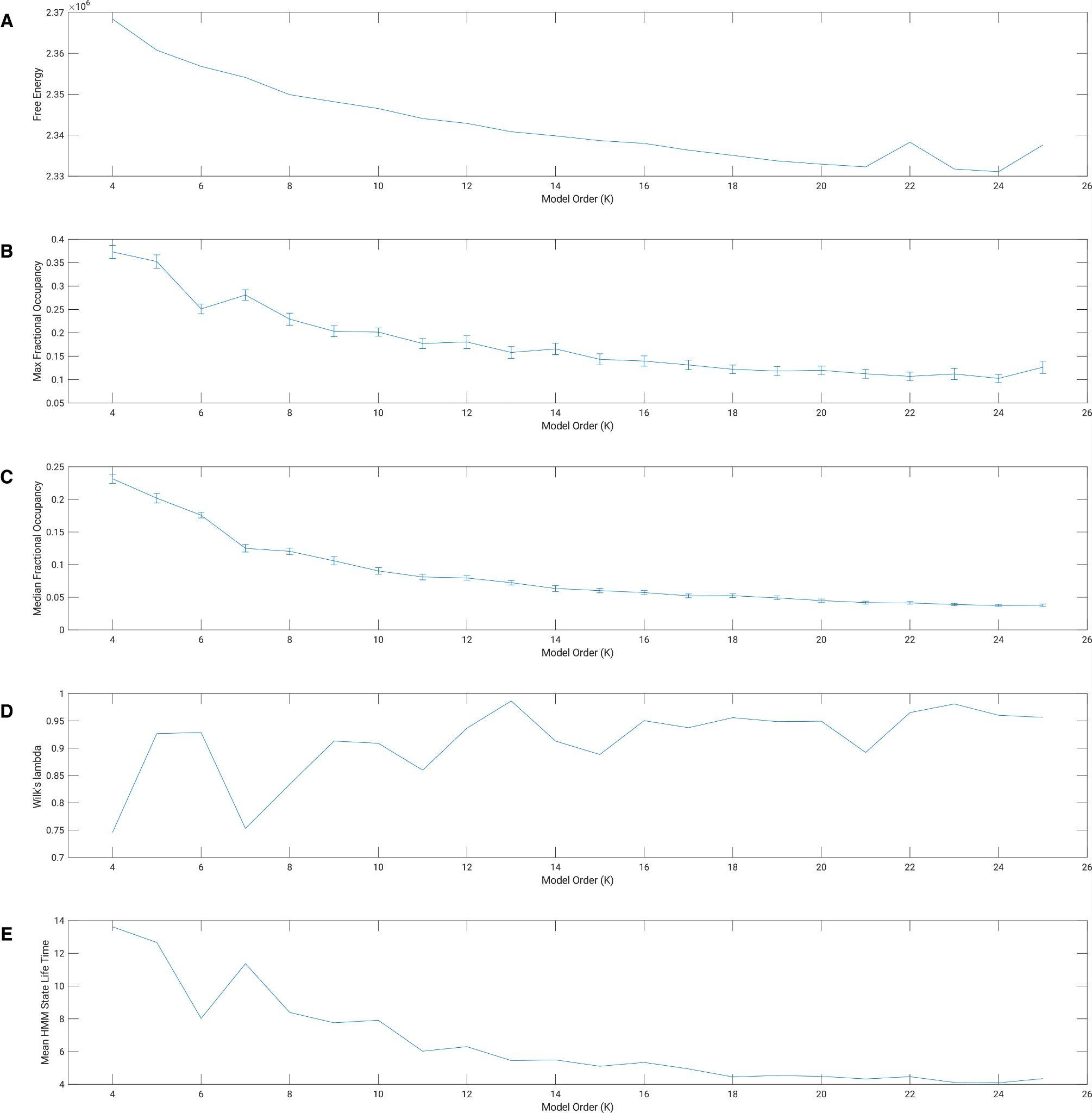

**Supplementary Figure 7.** Model evaluation parameters. The error bars represent the standard error of the mean. Panel **A**: free energy; Panel **B**: maximum Occupancy (percentage); Panel **C**: median Occupancy (percentage); Panel **D**: Wilk's Λ; Panel **E**: mean HMM state Lifetime (TR, 3-second).

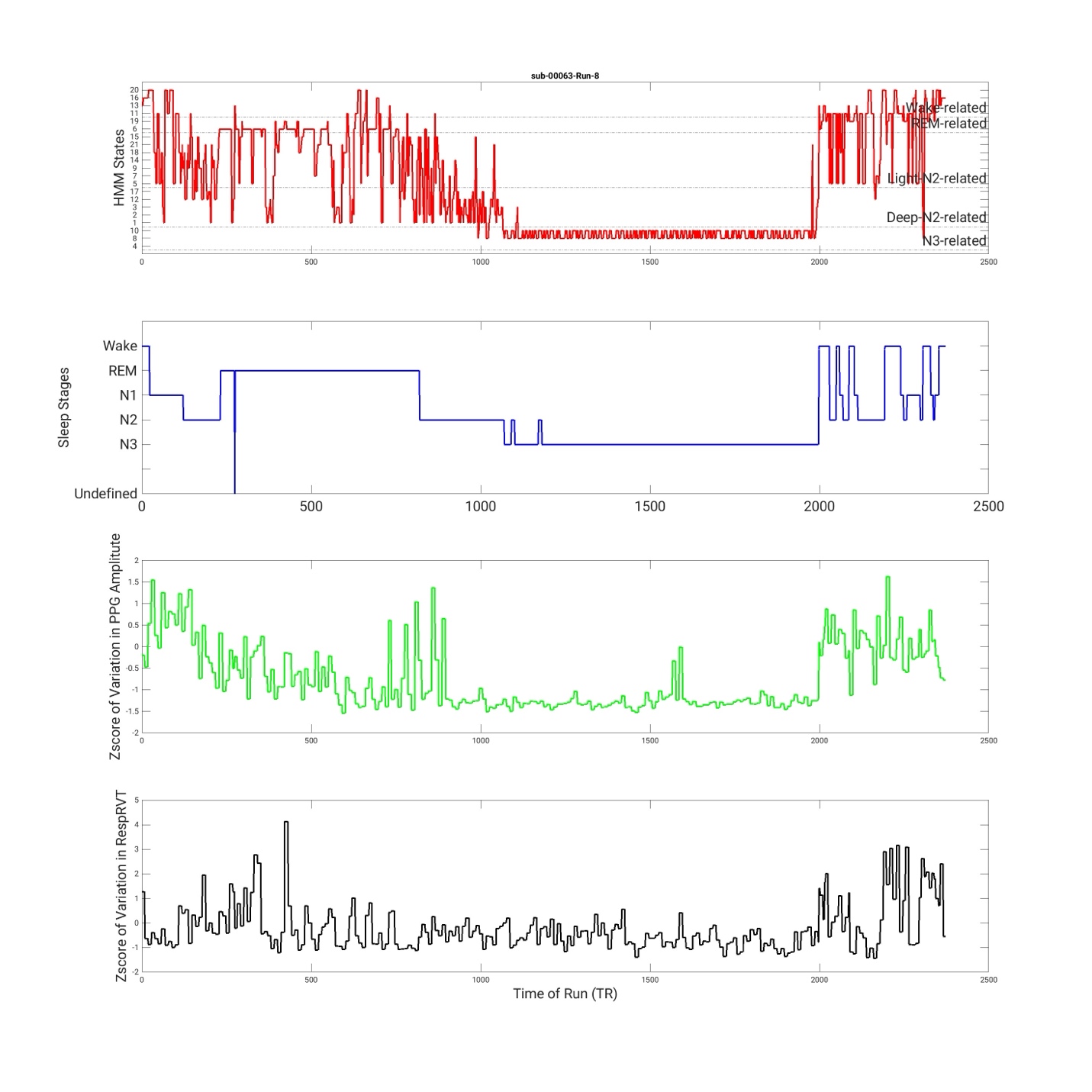

**Supplementary Figure 8.** State timecourse of HMM states and its associations with PSG stages, variation in PPG amplitude, and variations in RespRVT signals of an example run.

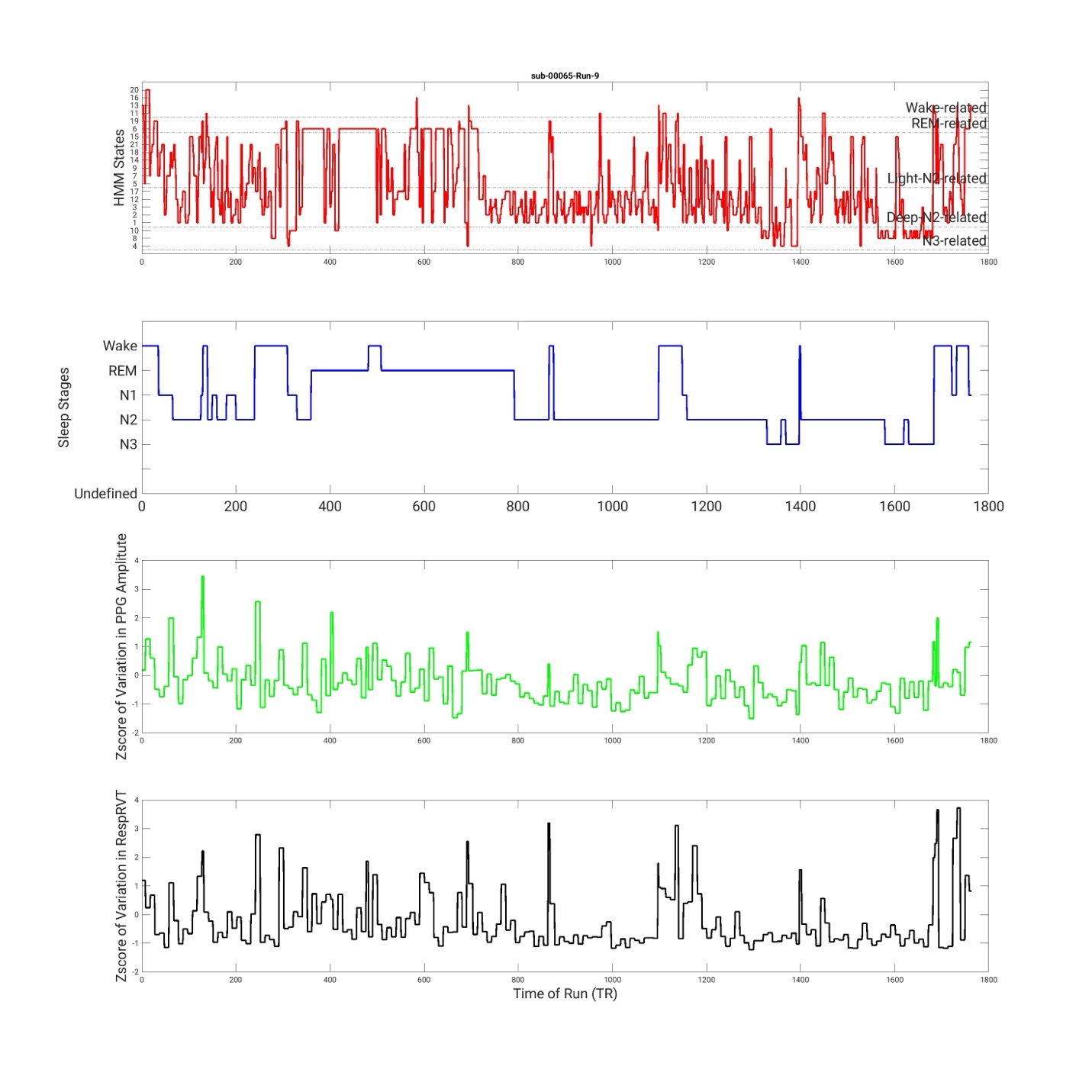

**Supplementary Figure 9.** State timecourse of HMM states and its associations with PSG stages, variation in PPG amplitude, and variations in RespRVT signals of a second example run.

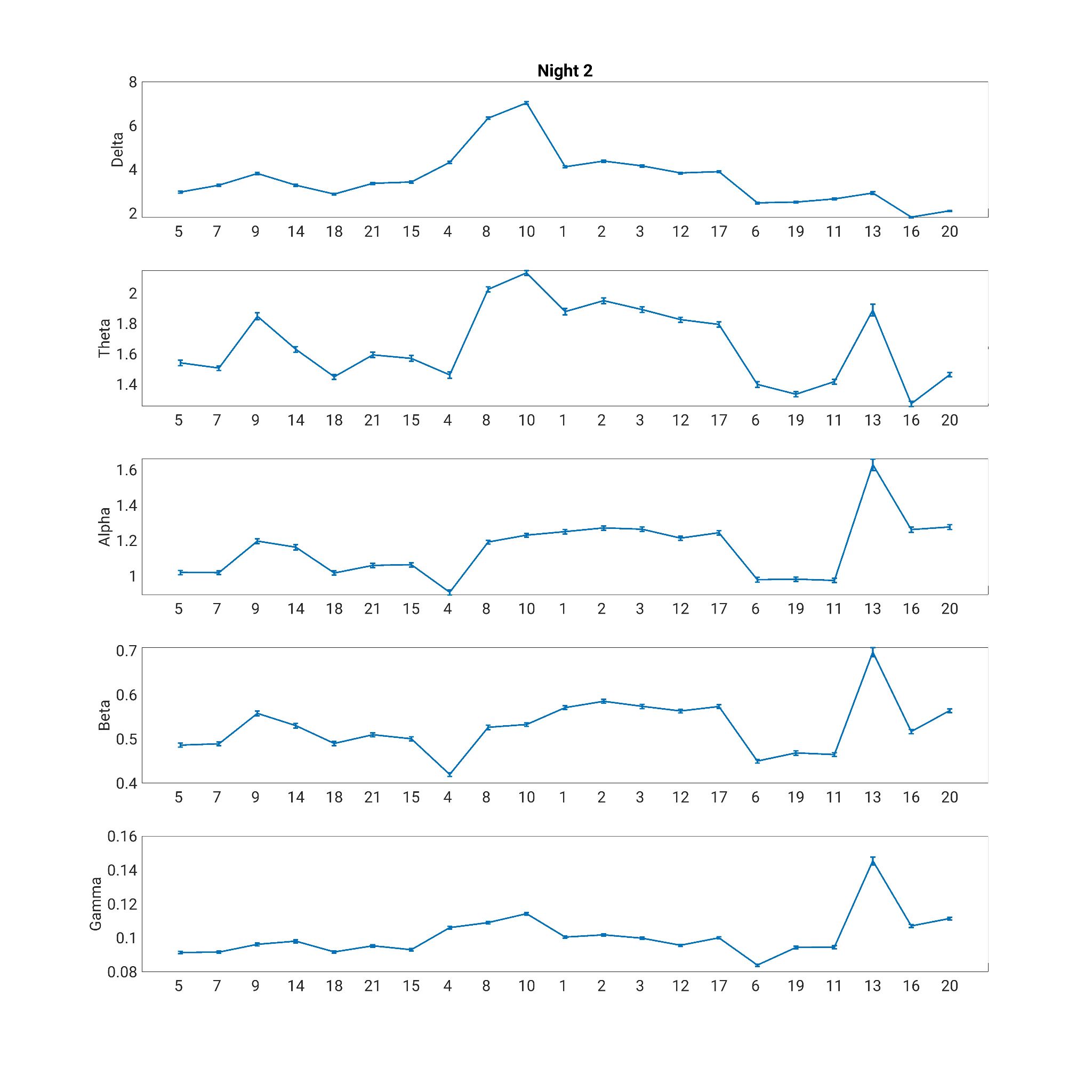

**Supplementary Figure 10.** EEG power spectrum associated with each HMM state during Night 2. The error bars represent the standard error of the mean.

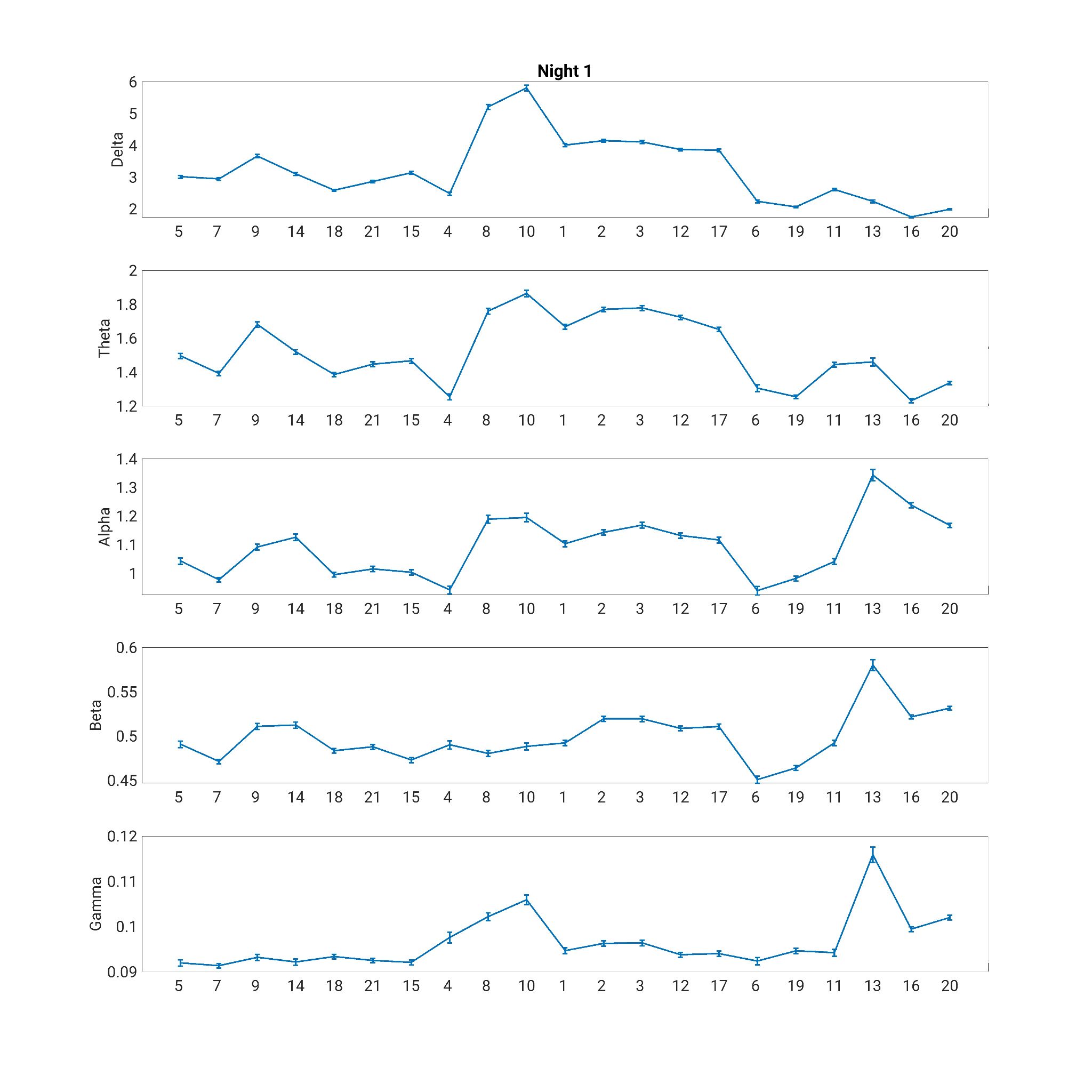

**Supplementary Figure 11.** EEG power spectrum associated with each HMM state during Night 1. The error bars represent the standard error of the mean.
